# White matter hyperintensity load varies depending on subjective cognitive decline criteria

**DOI:** 10.1101/2022.04.11.22273727

**Authors:** Cassandra Morrison, Mahsa Dadar, Sylvia Villeneuve, Simon Ducharme, D. Louis Collins, Alzheimer’s Disease Neuroimaging Initiative

## Abstract

**Background:** Increased age and cognitive impairment is associated with an increase in cerebrovascular pathology often measured as white matter hyperintensities (WMHs) on MRI. Whether WMH burden differs between cognitively unimpaired older adults with subjective cognitive decline (SCD+) and without subjective cognitive decline (SCD–) remains conflicting, and could be related to the methods used to identify SCD. Our goal was to examine if four common SCD classification methods are associated with different WMH accumulation patterns between SCD+ and SCD-.

**Methods:** A total of 535 cognitively unimpaired older adults with 1353 time points from the Alzheimer’s Disease Neuroimaging Initiative were included in this study. SCD was operationalized using four different methods: Cognitive Change Index (CCI), Everyday Cognition Scale (ECog), ECog+Worry, and Worry. Linear mixed-effects models were used to investigate the associations between SCD and overall and regional WMH burden.

**Results:** Overall temporal WMH burden differences were only observed with the Worry questionnaire. Higher WMH burden change over time was observed in SCD+ compared to SCD– in the temporal and parietal regions using the CCI (temporal, *p*=.01; parietal *p*=.03) and ECog (temporal, *p*=.03; parietal *p*=.01). For both the ECog+Worry and Worry questionnaire, change in WMH burden over time was increased in SCD+ compared to SCD- for overall, frontal, temporal, and parietal WMH burden (*p*<.05).

**Conclusion:** These results show that WMH burden differs between SCD+ and SCD– depending on the questionnaire and the approach (regional/global) used to measure WMHs. The various methods used to define SCD may reflect different types of underlying pathologies.

## Introduction

Subjective cognitive decline (SCD) is defined as self-reported deficits in cognitive functioning in the absence of objective cognitive decline [1]. People with SCD have increased risk of showing AD-related neurodegeneration and for being diagnosed with clinically probable Alzheimer’s disease (AD) [1–3]. The heightened risk for AD and increased neurodegeneration in cognitively unimpaired older adults with SCD (SCD+) compared to healthy older adults without SCD (SCD-) has led to the understanding that a subset of those who experience SCD are in the “preclinical AD” phase [1,2]. Nevertheless, many people who report SCD do not develop progressive cognitive decline. A meta-analysis found that in SCD+, only 27% progressed to mild cognitive impairment (MCI) and 14% progressed to dementia in a follow-up over 4 years, with an annual progression rate to MCI and dementia ranging from 2.3%-6.6% [4]. In addition to the varying progression rates, whether people with SCD have more neurodegeneration than healthy older adults remains relatively conflicting [5].

Varying conversion rates and neurodegeneration differences could be related to the numerous methodologies used to operationalize SCD. Previously, researchers observed that four commonly used methods to define SCD yielded distinct patterns of future cognitive decline and brain atrophy in cognitively unimpaired older adults [6]. While the cognitive change index (CCI) was associated with future cognitive decline as measured by the Alzheimer’s Disease Assessment scale-13, the Everyday Cognition Scale (ECog) was associated with cognitive decline as measured by the Montreal Cognitive Assessment. When examining atrophy differences, a relationship between hippocampal change and SCD was observed using the CCI and a Worry question. The ECog was not associated with hippocampal change but instead related to atrophy in superior temporal regions. These findings suggest that different SCD classification methods may measure different underlying pathologies leading to different types of cognitive decline.

A common pathological change in aging and cognitive decline results from small vessel cerebrovascular disease that appears as white matter hyperintensities (WMHs) in T2w and FLAIR MRI. WMHs often occur as a result of vascular risk factors such as obesity and high blood pressure [7]. Elevated WMH burden is associated with cognitive deterioration in healthy aging [8,9] and MCI and AD [10] as well as the risk of conversion to MCI and AD in healthy older adults [11–13]. Few studies have observed WMH differences between SCD+ and SCD-. When asking cognitively unimpaired older adults if they have changes in memory or thinking (regardless of worry), researchers observed no difference in WMHs in SCD+ vs. SCD- [14]. However, being concerned about changes in ones’ memory and thinking is a factor that, when endorsed, increases the risk of progression to dementia [1]. While one study of cognitively unimpaired older adults with worries regarding their memory complaints, SCD+ had more WMH load SCD- [15], another study found no WMH load SCD+ vs. SCD- differences [16]. The use of small sample sizes combined with different SCD questionnaires makes it difficult to compare WMH group differences between studies.

We hypothesize that subtle differences in pathological processes, impacting different areas of the brain, can lead to slightly different clinical symptoms that are captured by different SCD measures. The current study will provide insight into which SCD definition(s) is/are most associated with or most sensitive to vascular neuropathology, and if the four SCD definitions are associated with different pathological subtypes in WMH burden and WMH change over time. The goal of this study was to evaluate total and regional WMH burden to examine how they differ between the four SCD questionnaires.

## Methods

### Alzheimer’s Disease Neuroimaging Initiative

Data used in the preparation of this article were obtained from the Alzheimer’s Disease Neuroimaging Initiative (ADNI) database (adni.loni.usc.edu). The ADNI was launched in 2003 as a public-private partnership, led by Principal Investigator Michael W. Weiner, MD. The primary goal of ADNI has been to test whether serial MRI, positron emission tomography (PET), other biological markers, and clinical and neuropsychological assessment can be combined to measure the progression of mild cognitive impairment and early AD. The study received ethical approval from the review boards of all participating institutions. Written informed consent was obtained from participants or their study partner. Participants were selected only from ADNI-2 and ADNI-3 because those are the only two cohorts to complete questionnaires designed to measure subjective cognitive decline.

### Participants

Full inclusion and exclusion criteria are available at www.adni-info.org. Briefly, cognitively unimpaired older adults were between 55 and 90 at the time of recruitment, exhibiting no evidence of depression, no evidence of memory impairment as measured by the Wechsler Memory Scale, and no evidence of impaired global cognition as measured by the Mini Mental Status Examination (MMSE) or Clinical Dementia Rating (CDR). We used the same four SCD definitions as Morrison et al. [6] to classify SCD- and SCD+. The four groups were defined as:

- CCI: Participants were considered SCD+ if they had self-reported significant memory concern as quantified by a score of ≥ 16 on the first 12 items (representing memory changes) on the CCI. This score was selected based on previous research by Saykin et al. (2006) and because it is used by ADNI as a criterion to identify participants with significant memory concern [18].
- ECog: Participants were considered SCD+ if they endorsed any item on the ECog with a score ≥ 3. A score of ≥ 3 was used as it represents consistent SCD which has been shown to improve prognostic value of SCD for incident MCI [19].
- ECog + Worry: Participants were considered SCD+ if they had self-reported consistent SCD+ on any item from the ECog (again ≥ 3) as well as indicated worry/concern about their memory/thinking abilities.
- Worry: Participants were considered SCD+ if they indicated worry/ concern about their memory/thinking abilities, irrespective of their CCI or ECog scores.

A total of 619 cognitively normal older adults took part in ADNI-2 and ADNI-3. Of those, 535 had a total of 1353 MRI scans from which WMH measurements could be extracted from at least one timepoint and were used in the CCI and ECog analyses. Thirty people did not complete the question regarding worry about cognition therefore only 505 people with 1296 follow-ups were included in the ECog +Worry and Worry analyses. Follow-up periods ranged from 12 to 60 months.

Body mass index (BMI), systolic blood pressure, and diastolic blood pressure were downloaded from the ADNI public website and included as vascular risk factors for this study. BMI was calculated from height and weight information for the matching visit to the MRI scan.

### Structural MRI acquisition and processing

All participants were imaged using a 3T scanner with T1-weighted imaging parameters (see http://adni.loni.usc.edu/methods/mri-tool/mri-analysis/ for the detailed MRI acquisition protocol).

All longitudinal scans were downloaded from the ADNI website. T1w scans for each participant were pre-processed through our standard pipeline including noise reduction [20], intensity inhomogeneity correction [21], and intensity normalization into range [0-100]. The pre-processed images were then linearly (9 parameters: 3 translation, 3 rotation, and 3 scaling) [22] registered to the MNI-ICBM152-2009c average [23].

### WMH measurements

A previously validated WMH segmentation technique was employed to generate participant WMH measurements. This segmentation technique has been validated in multi-center studies such as the Parkinson’s Markers Initiative [24], National Alzheimer’s Coordinating Center [25], and importantly, has also been validated in ADNI [10] where a library of manual segmentations based on 50 ADNI participants (independent of those studied here) was created. WMHs were automatically segmented at baseline using the T1w contrasts, along with a set of location and intensity features obtained from a library of manually segmented scans in combination with a random forest classifier to detect the WMHs in new images [26,27]. WMH load was defined as the volume of all voxels as WMH in the standard space (in mm^3^) and are thus normalized for head size. The volumes of the WMHs for frontal, parietal, temporal, and occipital lobes as well as the entire brain were calculated based on regional masks from the Hammers atlas [26,28]. WMH volumes were log-transformed to achieve a more normal distribution. The quality of the registrations and WMH segmentations was visually verified by an experienced rater (author M.D.), blinded to diagnostic group. Only seven subjects did not pass this quality control step and were discarded, resulting in 535 people with 1353 follow-ups.

### Statistical Analysis

Analyses were performed using MATLAB R2019b. Independent sample t-tests and chi-square analysis were completed on demographic and clinical information. Baseline WMH load differences between SCD- and SCD+ were investigated using linear regressions and longitudinal data was investigated using linear mixed effects models to examine the association between WMH load (frontal, temporal, parietal, occipital, and total) and each SCD questionnaire. All results were corrected for multiple comparisons using false discovery rate (FDR), p-values are reported as raw values with significance then determined by FDR correction.

For baseline data, the categorical variable of interest was SCD group (i.e., SCD- vs SCD+) based on each questionnaire. The models also included sex, years of education, age at baseline (*Age_bl*), APOE4 status, body mass index (*BMI*), diastolic and systolic blood pressure (*BP_Diastolic* and *BP_Systolic*) as covariates.

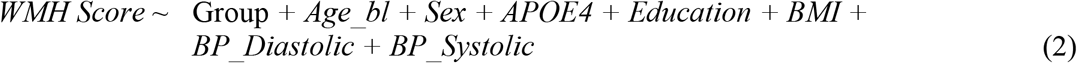

For longitudinal data, the categorical variable of interest was SCD group (i.e., SCD- vs SCD+) based on each questionnaire. The models also included time from baseline, sex, years of education, *Age_bl*, APOE4 status, *BMI, BP_Diastolic*, and *BP_Systolic* as covariates. Participant ID was included as a categorical random effect.

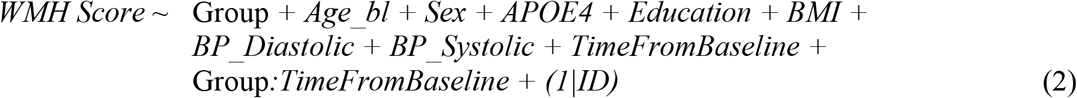

## Results

### Demographics

Table 1 presents demographic and clinical characteristics of study participants. For each of the four questionnaires, there were no significant SCD+ and SCD- group differences in any of the demographic or clinical features.

**Table 1:** Demographic and clinical characteristics for cognitively unimpaired older adults with and without subjective cognitive decline for the four questionnaires

| Demographic Information | CCI |  | ECOG |  | Ecog+Worry |  | Worry |  |
| --- | --- | --- | --- | --- | --- | --- | --- | --- |
|  | SCD-<br>n = 270 | SCD+<br>n = 265 | SCD-<br>n = 288 | SCD+<br>n = 247 | SCD-<br>n = 337 | SCD+<br>n = 168 | SCD-<br>n = 264 | SCD+<br>n = 241 |
| Age | 72.9 ± 6.6 | 72.1 ± 6.4 | 72.2 ± 6.4 | 73.4 ± 6.5 * | 72.2 ± 6.5 | 73.3 ± 6.2 | 72.3 ± 6.6 | 72.7 ± 6.3 |
| Education | 16.7 ± 2.4 | 16.8 ± 2.4 | 16.8 ± 2.5 | 16.6 ± 2.3 | 16.9 ± 2.4 | 16.4 ± 2.4 * | 17.01 ± 16.4 | 16.4 ± 2.5 * |
| AV-45 | 1.16 ± 0.3 | 1.12 ± 0.2 | 1.13 ± 0.2 | 1.14 ± 0.2 | 1.13 ± 0.2 | 1.15 ± 0.2 | 1.12 ± 0.2 | 1.16 ± 0.2 |
| APOE ε4+<sup>a</sup> | 77 (29%) | 91 (36%) | 74 (26%) | 74 (34%) | 105 (32%) | 54 (33%) | 72 (28%) | 87 (37%) * |
| Male sex | 122 (45%) | 103 (39%) | 119 (41%) | 106 (43%) | 151 (45%) | 64 (38%) | 120 (45%) | 95 (36%) |
| Diastolic BP | 74.3 ± 10.5 | 75.2 ± 8.3 | 74.9 ± 9.2 | 74.7 ± 9.8 | 75.1 ± 9.0 | 74.6 ± 10.1 | 75.2 ± 9.1 | 74.7 ± 9.6 |
| Systolic BP | 134.1 ± 18.9 | 135.2 ± 15.8 | 134.1 ± 17.3 | 135.2 ± 17.6 | 134.3 ± 16.7 | 135.1 ± 18.8 | 133.4 ± 16.4 | 135.8 ± 18.4 |
| BMI | 27.3 ± 5.2 | 27.7 ± 5.2 | 27.2 ± 5.3 | 27.8 ± 5.1 | 27.3 ± 5.2 | 27.8 ± 5.3 | 27.5 ± 5.4 | 27.5 ± 5.0 |

*Notes:* values are expressed as mean ± standard deviation, or number of participants (percentage %). \* = statistically significant differences between SCD- and SCD+ (at the p<0.05 level, uncorrected); however, none of these differences remain significant after correction for multiple comparisons. APOE ε4+ = number and percentage of people with at least one ε4 allele. <sup>a</sup> = APOE ε4+ status was missing for 15 participants. CCI = Cognitive Change Index. ECog = Everyday Cognition Scale. SCD- = Cognitively normal older adults without subjective cognitive decline. SCD+ = cognitively normal older adults with subjective cognitive decline. BMI = body mass index. BP = blood pressure.

### WMH baseline analyses

Table 2 summarizes the results of the linear regression models for all WMH regions and each questionnaire. For all questionnaires, increased age (*p*<.001) and diastolic blood pressure (*p*<.05) was associated with increased WMH burden except in the occipital region. There were no SCD+ vs. SCD- WMH differences using either the CCI or ECog questionnaires. SCD classification was associated with increased baseline WMH using the ECog+Worry questionnaire for total (*t*=2.47, *p*=.014), frontal (*t*=2.45, *p*=.015), and parietal (*t*=2.23, *p*=.026) regions compared to SCD-. SCD+ as classified using the worry questionnaire was associated with increased WMH burden compared to SCD- for total (*t*=2.15, *p*=.032), temporal (*t*=2.58, *p*=.01), and occipital (*t*=2.56 *p*=.011) regions.

**Table 2:** Linear regression model outputs showing differences baseline differences between cognitively unimpaired older adults with (SCD+) and without subjective cognitive decline (SCD-)

|  | CCI | ECog | ECog+Worry | Worry |
| --- | --- | --- | --- | --- |
| *Total* |  |  |  |  |
| Age_bl | **$t=8.04, p<.001$** | **$t=7.91, p<.001$** | **$t=7.92, p<.001$** | **$t=8.07, p<.001$** |
| BMI | $t=-0.20, p=.84$ | $t=-0.29, p=.77$ | $t=-0.33, p=.74$ | $t=-0.21, p=.83$ |
| Systolic BP | $t=-0.09, p=.93$ | $t=-0.21, p=.84$ | $t=-0.31, p=.76$ | $t=-0.34, p=.73$ |
| Diastolic BP | **$t=2.23, p=.026$** | **$t=2.33, p=.020$** | **$t=2.32, p=.021$** | **$t=2.37, p=.018$** |
| SCD Classification | $t=-0.24, p=.81$ | $t=1.28, p=.20$ | **$t=2.47, p=.014$** | **$t=2.15, p=.032$** |
| *Frontal* |  |  |  |  |
| Age_bl | **$t=7.50, p<.001$** | **$t=7.34, p<.001$** | **$t=7.34, p<.001$** | **$t=7.49, p<.001$** |
| BMI | $t=-0.46, p=.65$ | $t=-0.55, p=.58$ | $t=-0.60, p=.55$ | $t=-0.47, p=.63$ |
| Systolic BP | $t=0.82, p=.41$ | $t=0.75, p=.45$ | $t=0.66, p=.51$ | $t=0.67, p=.50$ |
| Diastolic BP | $t=1.33, p=.19$ | $t=1.40, p=.16$ | $t=1.39, p=.17$ | $t=1.41, p=.16$ |
| SCD Classification | $t=0.26, p=.79$ | $t=1.25, p=.22$ | **$t=2.45, p=.015$** | $t=1.60, p=.11$ |
| *Temporal* |  |  |  |  |
| Age_bl | **$t=6.75, p<.001$** | **$t=6.67, p<.001$** | **$t=6.65, p<.001$** | **$t=6.77, p<.001$** |
| BMI | $t=-1.17, p=.24$ | $t=-1.22, p=.22$ | $t=-1.27, p=.20$ | $t=-1.18, p=.24$ |
| Systolic BP | $t=-0.85, p=.40$ | $t=-0.94, p=.35$ | $t=-1.03, p=.30$ | $t=-1.17, p=.24$ |
| Diastolic BP | **$t=2.76, p=.006$** | **$t=2.82, p=.004$** | **$t=2.83, p=.005$** | **$t=2.93, p=.003$** |
| SCD Classification | $t=-0.42, p=.67$ | $t=0.78, p=.44$ | $t=1.86, p=.063$ | **$t=2.58, p=.01$** |
| *Parietal* |  |  |  |  |
| Age_bl | **$t=8.26, p<.001$** | **$t=8.15, p<.001$** | **$t=8.18, p<.001$** | **$t=8.31, p<.001$** |
| BMI | $t=-0.10, p=.92$ | $t=-0.19, p=.85$ | $t=-0.22, p=.83$ | $t=-0.10, p=.92$ |
| Systolic BP | $t=-0.66, p=.51$ | $t=-0.86, p=.39$ | $t=-0.93, p=.35$ | $t=-0.95, p=.34$ |
| Diastolic BP | **$t=2.66, p=.008$** | **$t=2.78, p=.006$** | **$t=2.76, p=.006$** | **$t=2.80, p=.005$** |
| SCD Classification | $t=-1.04, p=.30$ | $t=1.36, p=.17$ | **$t=2.23, p=.026$** | $t=1.85, p=.066$ |
| *Occipital* |  |  |  |  |
| Age_bl | $t=1.83, p=.07$ | $t=1.79, p=.07$ | $t=1.75, p=.08$ | $t=1.80, p=.40$ |
| BMI | $t=1.60, p=.11$ | $t=1.57, p=.12$ | $t=-1.53, p=.13$ | $t=1.59, p=.11$ |
| Systolic BP | $t=-0.12, p=.91$ | $t=-0.15, p=.88$ | $t=-0.22, p=.83$ | $t=-0.40, p=.69$ |
| Diastolic BP | $t=1.31, p=.19$ | $t=1.34, p=.18$ | $t=1.35, p=.18$ | $t=1.47, p=.14$ |
| SCD Classification | $t=-0.06, p=.95$ | $t=0.35, p=.72$ | $t=1.20, p=.23$ | **$t=2.56 p=.011$** |

*Notes:* CCI = Cognitive Change Index. ECog = Everyday Cognition Scale. BMI = body mass index, BP = blood pressure. Bolded values represent those that remained significant after correction for multiple comparisons using FDR.

### WMH longitudinal analyses

Table 3 summarizes the linear mixed effects model results for all WMH regions by questionnaire. Figure 1 presents the longitudinal change in WMH by follow-up duration for each group and region. For all questionnaires, baseline age and time from baseline had significant associations with frontal, temporal, parietal, occipital, and total WMH load. That is, with increased age at baseline and time from baseline, WMH load was higher for all regions and all questionnaires.

**Table 3:** Linear mixed effects model outputs showing longitudinal differences between cognitively unimpaired older adults with (SCD+) and without subjective cognitive decline (SCD-)

|  | CCI | ECog | ECog+Worry | Worry |
| --- | --- | --- | --- | --- |
| *Total* |  |  |  |  |
| Age_bl | **$t=9.19, p<.001$** | **$t=9.05, p<.001$** | **$t=9.00, p<.001$** | **$t=9.10, p<.001$** |
| TimeFromBaseline | **$t=18.35, p<.001$** | **$t=14.38, p<.001$** | **$t=15.88, p<.001$** | **$t=13.95, p<.001$** |
| BMI | $t=0.71, p=.48$ | $t=0.69, p=.48$ | $t=0.44, p=.66$ | $t=0.47, p=.64$ |
| Systolic BP | **$t=2.21, p=.028$** | **$t=2.21, p=.027$** | $t=2.06, p=.039^*$ | $t=2.07, p=.038^*$ |
| Diastolic BP | $t=0.19, p=.85$ | $t=0.09, p=.92$ | $t=0.12, p=.91$ | $t=0.14, p=.88$ |
| SCD Classification | $t=-0.22, p=.82$ | $t=0.77, p=.44$ | $t=1.97, p=.048^*$ | $t=1.83, p=.067$ |
| SCD:TimeFromBaseline | $t=1.03, p=.30$ | $t=1.63, p=.10$ | **$t=3.80, p=.001$** | **$t=3.86, p<.001$** |
| *Frontal* |  |  |  |  |
| Age_bl | **$t=8.93, p<.001$** | **$t=8.79, p<.001$** | **$t=8.76, p<.001$** | **$t=8.85, p<.001$** |
| TimeFromBaseline | **$t=16.81, p<.001$** | **$t=13.47, p<.001$** | **$t=14.04, p<.001$** | **$t=11.96, p<.001$** |
| BMI | $t=1.33, p=.18$ | $t=1.31, p=.19$ | $t=1.01, p=.31$ | $t=1.01, p=.31$ |
| Systolic BP | **$t=2.72, p=.007$** | **$t=2.73, p=.007$** | **$t=2.58, p=.01$** | **$t=2.61, p=.008$** |
| Diastolic BP | $t=-0.48, p=.63$ | $t=-0.52, p=.60$ | $t=-0.37, p=.71$ | $t=-0.37, p=.71$ |
| SCD Classification | $t=0.29, p=.78$ | $t=0.73, p=.47$ | $t=2.01, p=.044^*$ | $t=1.41, p=.16$ |
| SCD:TimeFromBaseline | $t=0.34, p=.73$ | $t=0.69, p=.49$ | **$t=3.48, p<.001$** | **$t=4.08, p<.001$** |
| *Temporal* |  |  |  |  |
| Age_bl | **$t=7.60, p<.001$** | **$t=7.50, p<.001$** | **$t=7.44, p<.001$** | **$t=7.50, p<.001$** |
| TimeFromBaseline | **$t=12.73, p<.001$** | **$t=10.02, p<.001$** | **$t=11.82, p<.001$** | **$t=10.49, p<.001$** |
| BMI | $t=-0.34, p=.73$ | $t=-0.27, p=.79$ | $t=-0.43, p=.67$ | $t=-0.40, p=.69$ |
| Systolic BP | $t=0.48, p=.63$ | $t=0.43, p=.66$ | $t=0.35, p=.72$ | $t=0.33, p=.74$ |
| Diastolic BP | $t=1.16, p=.24$ | $t=1.04, p=.30$ | $t=0.86, p=.39$ | $t=0.90, p=.37$ |
| SCD Classification | $t=-0.34, p=.73$ | $t=0.43, p=.79$ | $t=1.47, p=.14$ | **$t=2.19, p=.028$** |
| SCD:TimeFromBaseline | **$t=2.56, p=.01$** | **$t=2.23, p=.03$** | **$t=3.28, p=.001$** | **$t=3.06, p=.002$** |
| *Parietal* |  |  |  |  |
| Age_bl | **$t=8.90, p<.001$** | **$t=8.83, p<.001$** | **$t=8.82, p<.001$** | **$t=8.91, p<.001$** |
| TimeFromBaseline | **$t=17.09, p<.001$** | **$t=13.27, p<.001$** | **$t=15.81, p<.001$** | **$t=14.34, p<.001$** |
| BMI | $t=0.19, p=.85$ | $t=0.20, p=.84$ | $t=0.04, p=.97$ | $t=0.12, p=.91$ |
| Systolic BP | $t=0.63, p=.53$ | $t=0.59, p=.55$ | $t=0.56, p=.58$ | $t=0.55, p=.58$ |
| Diastolic BP | $t=0.73, p=.47$ | $t=0.58, p=.56$ | $t=0.59, p=.55$ | $t=0.63, p=.53$ |
| SCD Classification | $t=-1.16, p=.25$ | $t=1.07, p=.28$ | $t=1.93, p=.05^*$ | $t=1.72, p=.085$ |
| SCD:TimeFromBaseline | **$t=2.23, p=.03$** | **$t=2.50, p=.01$** | **$t=2.95, p=.003$** | **$t=2.45, p=.01$** |
| *Occipital* |  |  |  |  |
| Age_bl | **$t=2.61, p=.009$** | **$t=2.58, p=.009$** | **$t=2.51, p=.01$** | **$t=2.49, p=.01$** |
| TimeFromBaseline | **$t=7.17, p<.001$** | **$t=5.22, p<.001$** | **$t=5.77, p<.001$** | **$t=4.89, p<.001$** |
| BMI | $t=-0.51, p=.61$ | $t=-0.56, p=.57$ | $t=-0.56, p=.57$ | $t=-0.58, p=.56$ |
| Systolic BP | $t=1.99, p=.047^*$ | $t=2.02, p=.044^*$ | $t=1.99, p=.046^*$ | $t=1.97, p=.049^*$ |
| Diastolic BP | $t=-0.18, p=.86$ | $t=-0.21, p=.83$ | $t=-0.37, p=.71$ | $t=-0.36, p=.72$ |
| SCD Classification | $t=0.01, p=.99$ | $t=0.09, p=.93$ | $t=0.66, p=.51$ | $t=1.95, p=.051$ |
| SCD:TimeFromBaseline | $t=-0.68, p=.50$ | $t=0.47, p=.64$ | $t=1.41, p=.16$ | $t=1.67, p=.094$ |

Notes: CCI = Cognitive Change Index. ECog = Everyday Cognition Scale. BMI = body mass index, BP = blood pressure.
\* Represents results that are significant ( $p<0.05$ , uncorrected) and bolded values represent those that remained significant after correction for multiple comparisons using FDR.

**Figure 1:**
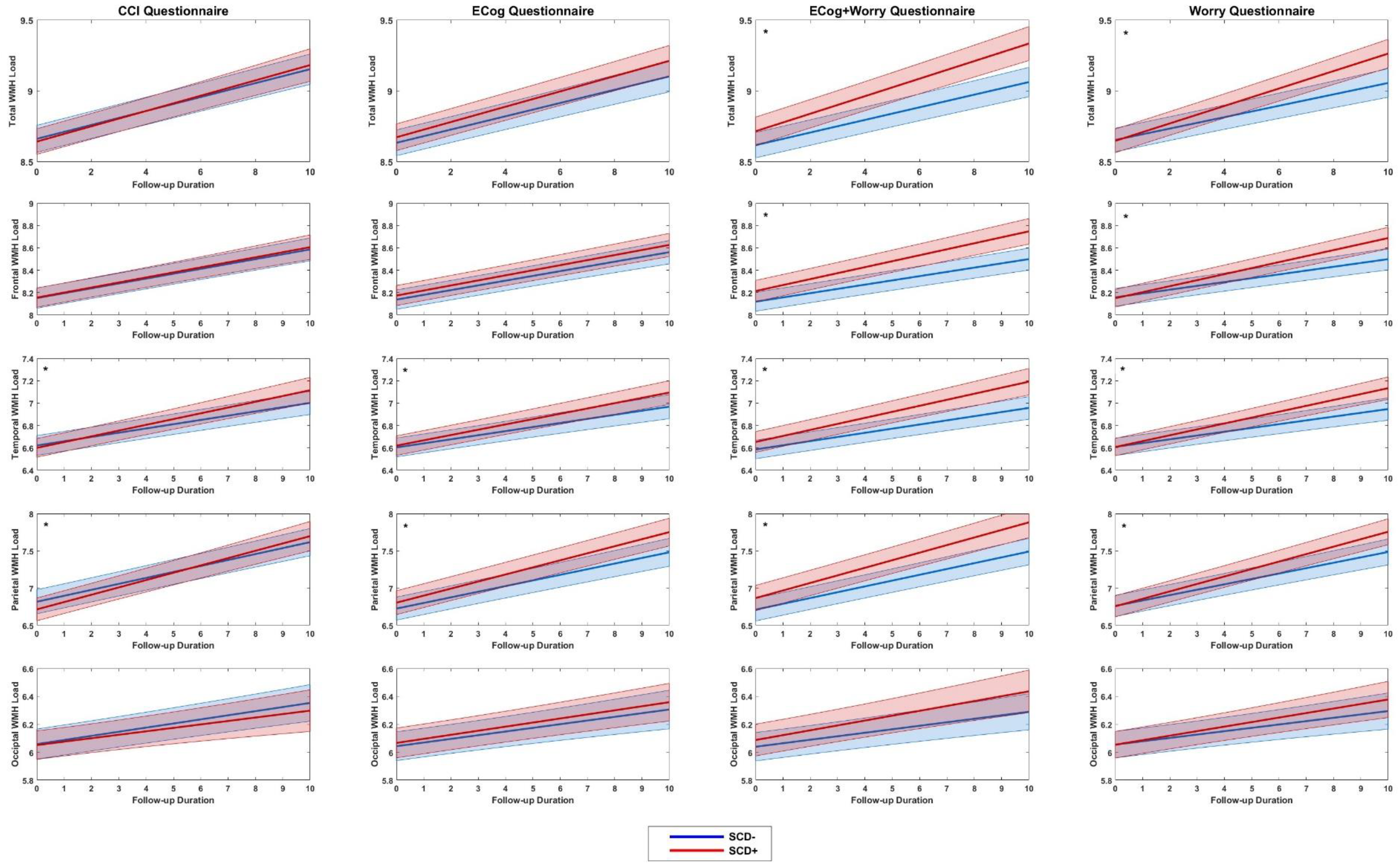
Linear mixed effects models showing longitudinal WMH change over time for SCD+ and SCD-. Notes: SCD+ = cognitively unimpaired older adult with subjective cognitive decline. SCD- = cognitively unimpaired older adult without subjective cognitive decline. WMH = white matter hyperintesnity. CCI= cognitive change index. ECog = Everyday Cognition Scale. * represents figures for which WMH rate of change over time differed between the groups.

For vascular risk factors, increased systolic blood pressure was associated with increased total WMH load only for CCI and ECog questionnaires. This association was also found in the frontal lobes for all four SCD questionnaires (*p*<.01). No associations were found for BMI or diastolic BP that survived corrections for multiple comparisons.

For the CCI questionnaire, there were no main effects of SCD diagnosis on total or regional WMH volume. The interaction between SCD classifcation and time from baseline on WMH load was significant in the temporal (*t*=2.56, *p*=.01) and parietal (*t*=2.23, *p*=.03) regions. Using the ECog, no main effect of SCD diagnosis on total or regional WMH volume. Significant interactions between SCD diagnosis and time from baseline were observed at the temporal *(t*=2.23, *p*=.03) and parietal (*t*=2.50, *p*=.01) regions. These findings indicate that for the CCI and ECog questionnaires, that rate of change in WMH load over time was increased for SCD+ vs. SCD- in both temporal and parietal regions.

Using the ECog+Worry questionnaire, no main effects of SCD diagnosis on total or regional WMH volume remained significant after correction for multiple comparisons. The interaction between SCD diagnosis and time from baseline was significant for frontal (*t*=3.48, *p*<.001), temporal (*t*=3.28, *p*=.001), parietal (*t*=2.95, *p*=.003), and total (*t*=3.80 *p*=.001), but not occipital (*t*=1.41, *p*=.16) WMH load. The Worry question showed a significant main effect for SCD diagnosis on WMH load in only the temporal region (*t=*2.19, *p*=.028). Interactions between SCD diagnosis and time from baseline and WMH load were significant for frontal (*t*=4.08, *p*<.001), temporal (*t*=3.06, *p*=.002), parietal (*t*=2.45, *p*=.01) and total (*t*=3.86 *p*<.001). These findings indicated that for ECog+Worry and Worry SCD questionnaires, the rate of change in WMH load over time was increased for SCD+ vs. SCD- for all regions except occipital.

## Discussion

Previous research has observed that increased WMH burden is associated with cognitive decline and conversion to dementia (Bangen et al., 2018; Dadar et al., 2019; Prins et al., 2004). SCD has also been associated with future cognitive decline, increased conversion to dementia [1,3], and increased atrophy compared to healthy older adults [6]. However, these associations depend on the questionnaires used to classify SCD. The relationship between SCD and WMHs remains relatively unexplored. The current study compared WMH load in SCD+ vs SCD- using four commonly used methods of classifying SCD, with the goal of examining if the four methods were associated with different patterns of vascular pathology. Our findings indicate that the four SCD questionnaires are associated with different patterns of WMH accumulation.

Similar results were obtained for both CCI and ECog definitions of SCD. The increase of WMH load over time in the SCD+ participants was greater compared to the SCD- participants in the temporal and parietal regions. Previous research examining WMH differences in people with amnestic MCI, who are likely to develop AD, have shown increases in WMHs in temporal and occipital regions [29]. Increased WMH volume in the parietal lobe has also been shown to predict incident AD in healthy older adults [30]. In a post-mortem studying examining white matter lesions (WMLs), parietal WMLs appear to be most associated with AD-associated degenerative mechanisms, whereas frontal WMLs are associated with both AD-associated degenerative pathology and small vessel disease associated mechanisms [31]. Taken together with past research, our findings suggest that both the CCI and ECog questionnaire may be associated with WMH burden that is observed in people with AD. A previous study found that using ECog to define SCD+ was associated with atrophy in the superior temporal region. On the other hand, using CCI to define SCD+ was associated with hippocampal change [6], an area known to be associated with early AD pathology. Taken together, the CCI definition of SCD+ is associated with both WMH burden in the temporal and parietal regions and hippocampal change, suggesting that this definition may be more sensitive to a future AD diagnosis than the ECog definition of SCD+. Of course, when applied in clinical settings an AD biomarker positivity is needed to confirm the diagnosis.

Differences in total and frontal WMH burden change over time were observed using the ECog+Worry questionnaire. These differences were likely driven by the Worry definition, as observed by ECog having no association SCD classification in WMH total or the frontal region, whereas Worry was associated with group differences for both total frontal as well as the temporal region WMH burden. These findings suggest that SCD+ as classified by Worry is associated with a more widespread regional vascular pathology than SCD+ as classified by either the ECog or CCI. Previous research has observed that non-amnestic MCI is associated with a widespread WMH accumulation (frontal, parietal, temporal, and occipital), whereas amnestic MCI is associated with elevated WMH burden in the temporal and occipital areas relative to normal controls [29]. It is thus possible that the Worry questionnaire is detecting individuals who may develop non-amnestic MCI. Future research is needed to examine conversion rates in these individuals and confirm this hypothesis.

Depending on SCD classification method and whether regional or total WMH burden was measured, our results here replicate both the findings that observe SCD+ vs. SCD- WMH group differences [15,17] and those that do not [14,16]. The limitation in these past studies is that they examined only baseline WMH volume as opposed to longitudinal change, which could explain the lack of group differences observed in several of these papers. In our study, only SCD+ defined using Worry and ECog+Worry was associated with increased baseline WMHs. However, all definitions were associated with increased change in WMH burden over time in SCD+. This finding follows previous research suggesting that increased WMHs may occur before symptom onset [32]. The large sample size in this study has sufficient power to conclude that subtle WMH differences exist between SCD+ and SCD-, and that these differences are most prominent when examining change over time.

Many studies examining WMH differences in SCD measure overall WMH burden [14–16] as opposed to regional WMHs [17]. The current findings suggest that examining regional differences will influence whether higher WMH burden is observed in SCD+ compared to SCD-. When examining total WMH burden change over time, both the CCI and ECog definitions show no differences between SCD+ vs. SCD-. However, when examining WMHs in a regional approach, group differences are found in the temporal and parietal regions. WMH change over time because of SCD+ diagnosis in both the temporal and parietal region is subtle and thus may be “washed out” when WMHs are examined in a whole brain metric. Future research should use regional approaches when examining WMH differences in healthy older adults.

There are a few limitations of the current study that should be noted. ADNI participants are highly educated, limiting generalizability to more representative samples. Although we examined change over time, additional research with longer follow-ups would improve our understanding of how different topographical distributions of WMH burden are associated with conversion to dementia. All ADNI protocols excluded individuals with a Hachinski score of greater than 4, thus associations reported here may be underestimated.

This study shows that change in WMH burden observed in SCD+ depends on both the definition used to define SCD and whether a regional or whole brain approach is used to measure WMH burden. Neuropathological changes in SCD may be too subtle to observe in a whole brain approach. Therefore, future research should examine WMH changes in SCD+ populations using a regional approach to accurately examine pathological changes. While the CCI and ECog questionnaires are associated with temporal and parietal WMH burden, the Worry definition is associated with a more widespread WMH accumulation in total, frontal, temporal, and parietal regions. That is, cognitively unimpaired older adults who endorse being worried about their memory functioning will have the most WMH pathology compared to the other definitions. Based on these findings, the four different SCD questionnaires are associated with different WMH accumulation patterns.

## Data Availability

Data is available from the ADNI online database at : http://www.adni-info.org/

## Acknowledgments

Data collection and sharing for this project was funded by the Alzheimer’s Disease Neuroimaging Initiative (ADNI) (National Institutes of Health Grant U01 AG024904) and DOD ADNI (Department of Defense award number W81XWH-12-2-0012). ADNI is funded by the National Institute on Aging, the National Institute of Biomedical Imaging and Bioengineering, and through generous contributions from the following: AbbVie, Alzheimer’s Association; Alzheimer’s Drug Discovery Foundation; Araclon Biotech; BioClinica, Inc.; Biogen; Bristol-Myers Squibb Company; CereSpir, Inc.; Cogstate; Eisai Inc.; Elan Pharmaceuticals, Inc.; Eli Lilly and Company; EuroImmun; F. Hoffmann-La Roche Ltd and its affiliated company Genentech, Inc.; Fujirebio; GE Healthcare; IXICO Ltd.; Janssen Alzheimer Immunotherapy Research & Development, LLC.; Johnson & Johnson Pharmaceutical Research & Development LLC.; Lumosity; Lundbeck; Merck & Co., Inc.; Meso Scale Diagnostics, LLC.; NeuroRx Research; Neurotrack Technologies; Novartis Pharmaceuticals Corporation; Pfizer Inc.; Piramal Imaging; Servier; Takeda Pharmaceutical Company; and Transition Therapeutics. The Canadian Institutes of Health Research is providing funds to support ADNI clinical sites in Canada. Private sector contributions are facilitated by the Foundation for the National Institutes of Health (www.fnih.org). The grantee organization is the Northern California Institute for Research and Education, and the study is coordinated by the Alzheimer’s Therapeutic Research Institute at the University of Southern California. ADNI data are disseminated by the Laboratory for Neuro Imaging at the University of Southern California.

